# An expert judgment model to predict early stages of the COVID-19 outbreak in the United States

**DOI:** 10.1101/2020.09.21.20196725

**Authors:** Thomas McAndrew, Nicholas G. Reich

## Abstract

During early stages of the COVID-19 pandemic, forecasts provided actionable information about disease transmission to public health decision-makers. Between February and May 2020, experts in infectious disease modeling made weekly predictions about the impact of the pandemic in the U.S. We aggregated these predictions into consensus predictions. In March and April 2020, experts predicted that the number of COVID-19 related deaths in the U.S. by the end of 2020 would be in the range of 150,000 to 250,000, with scenarios of near 1m deaths considered plausible. The wide range of possible future outcomes underscored the uncertainty surrounding the outbreak’s trajectory. Experts’ predictions of measurable short-term outcomes had varying levels of accuracy over the surveys but showed appropriate levels of uncertainty when aggregated. An expert consensus model can provide important insight early on in an emerging global catastrophe.

**One Sentence Summary:** Expert predictions provided valuable insight of future societal burden during the early stages of the COVID-19 pandemic in the US.

## Main Text

The first recorded world-wide COVID-19 cases were reported in December of 2019 [1], the World Health Organization declared the outbreak a Public Health Emergency of International Concern on January 30^th^, 2020, and on March 11^th^, 2020, after the virus began spreading to other continents [2], [3], the World Health Organization designated the outbreak a pandemic [4]. After the first COVID-19 case in the United States without known origin occurred in California in late February [5], a large-scale national effort aimed to prevent the spread of the disease.

As with previous outbreaks of other diseases [6]–[8], forecasts from computational models [9]– [12] assisted in planning and outbreak response in the first few months of the pandemic. However, these models faced three important challenges: a lack of reliable surveillance data, a paucity of data on models to explain SARS-COV-2 transmission dynamics, and unknown interactions between forecasts and future policy responses to the pandemic. As a result of these challenges in this phase, some models used by decision-makers faced criticism for a lack of accuracy [13].

Before the first US case of COVID-19, we aggregated probabilistic predictions weekly from experts in the modeling of infectious disease to support public health decision making [14].

Aggregating human predictions has shown positive results in many domains from ecology to economics [15]–[20]. In the context of infectious disease, human judgment (from a knowledgeable but not exclusively “expert” panel) produced accurate forecasts of seasonal influenza outbreaks in recent seasons [6]. In the early stages of a pandemic there is a tremendous amount of objective scientific work and subjective media attention that makes identifying key information difficult, but past work shows that experts familiar with the subject matter may be able to wade through this information and extract important relationships between data and forecasting targets [21]. Past work has shown that expert predictions are often well-calibrated, and crowdsourcing research suggests a non-expert crowd can accurately predict targets sensitive to societal change and governmental intervention [18]. This may be particularly important in the context of an emerging pandemic with fast-changing government responses. Subjective forecasts from individual experts are often well calibrated, but less accurate than forecasts from computational models [22]. But gains in accuracy have been found when individual human predictions are combined into a consensus prediction [15], [23], [24].

Between February 18^th^, 2020 and May 11^th^, 2020, we conducted thirteen weekly surveys of experts in the modeling of infectious disease [14]. Across these surveys, we asked 75 questions (40 with measurable outcomes) focused on the outbreak in the United States. A total of 41 experts contributed predictions, with an average of 18.6 responses each week (range: 15-22). We combined predictions into consensus distributions (see Methods). The survey results were released publicly every week and delivered directly to decision makers at state and federal health agencies. Experts responded to questions on a variety of topics including short- and long-term predictions of COVID-19 cases, hospitalizations, and deaths. Here we present results from questions that other computational models have tried to predict: the number of deaths due to COVID-19 in the US by the end of 2020, the total number of SARS-COV-2 infections in the US, and the number of confirmed cases one week ahead. Expert consensus forecasts for all questions are stored in a public GitHub repository [14].

Across five surveys administered in March, April, and May, we asked experts to predict the number of COVID-19 deaths in the US by the end of 2020. The consensus median ranged from 150,000 to more than 250,000 (Fig. 1) corresponding to between 4 and 7 times the average number of annual deaths in the US due to seasonal influenza [25]. There was considerable uncertainty around these predictions: the lower bound of the five 90% prediction intervals ranged from 6,000 (on March 16^th^) to 118,000 (on May 5^th^), and the upper bounds ranged from 517,000 (on April 20^th^) to 1,700,000 (on March 30^th^). As of Aug. 26, 2020, over 171,000 cumulative deaths due to COVID-19 were reported in the US.

**Fig. 1.**
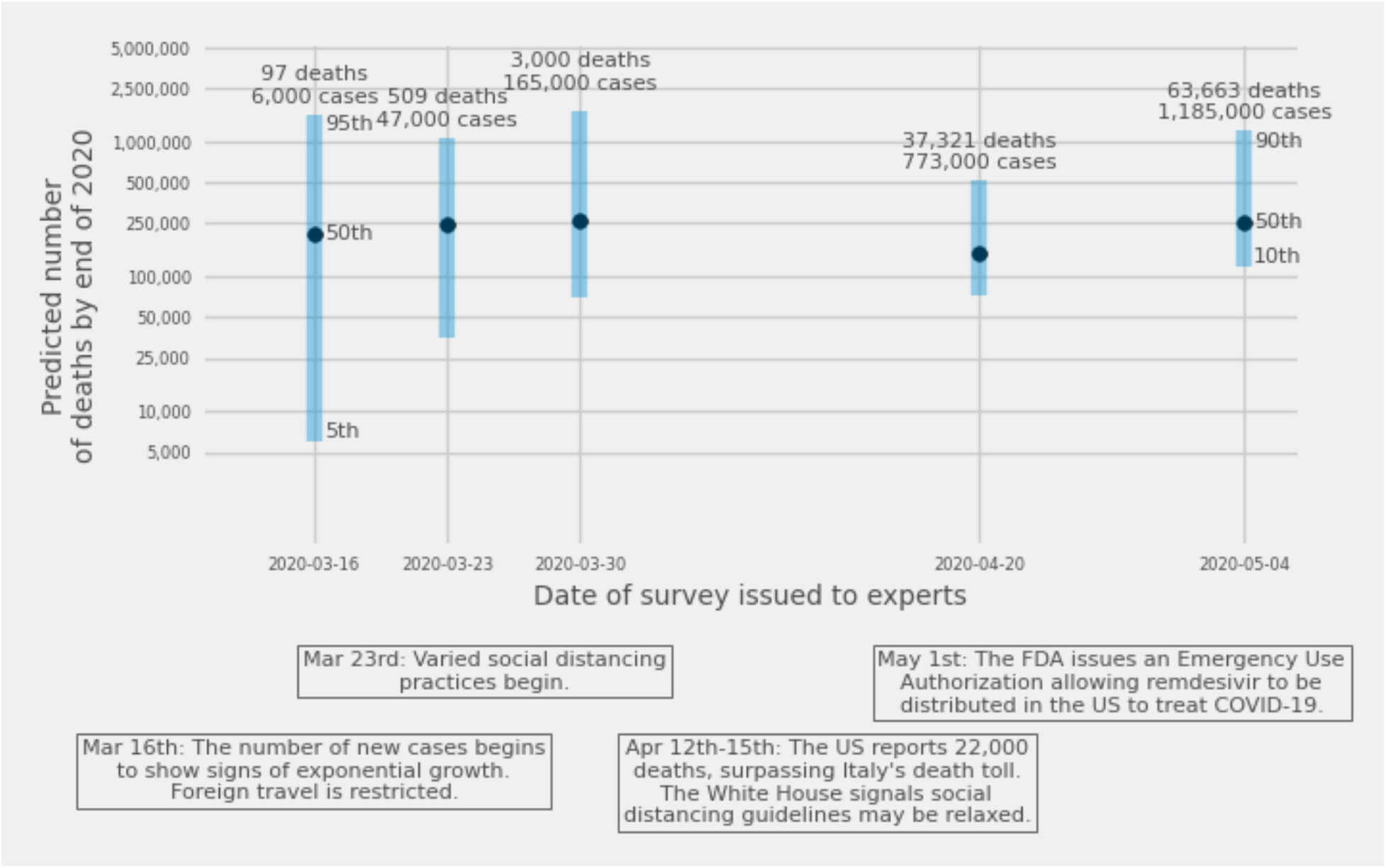
Expert consensus predictions of the total number of deaths by the end of 2020, from five surveys asked between March 16 and May 4, 2020. Points show the median estimate. Bars show 90% prediction intervals for the first four surveys and an 80% prediction interval for the fifth survey. The first three surveys shown above asked experts for predictions of the smallest, most likely, and largest number of deaths; the fourth survey asked for 5^th^, 50^th^, and 95^th^ predictive percentiles, and the fifth survey asked for 10th, 50th, and 90th percentiles (see Methods). The counts of cases and deaths above each prediction are the numbers reported by Covidtracker.com on the date each survey was issued, and the text below the x-axis provides context of national headlines during the times the surveys were open to responses.

**Fig. 2.**
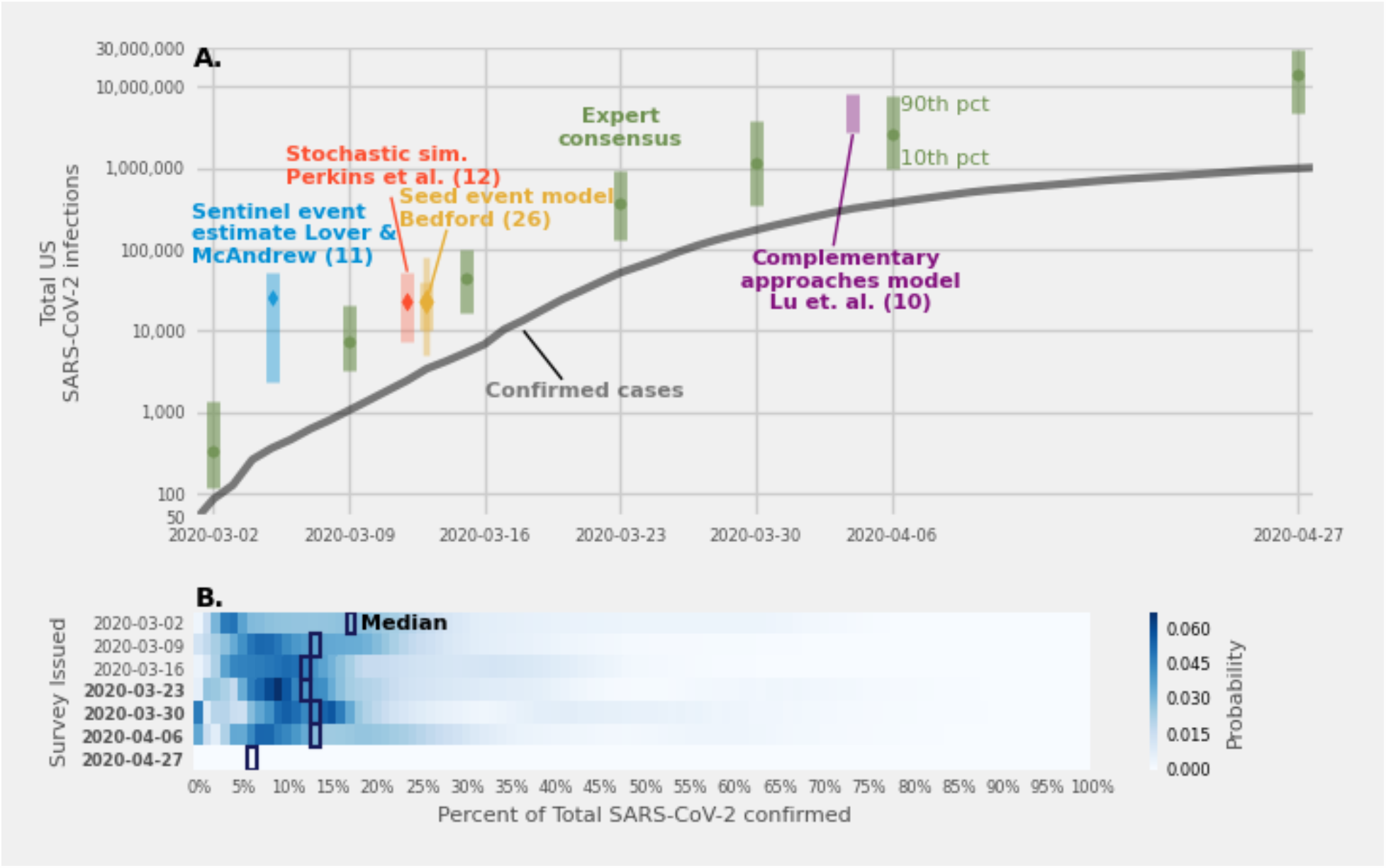
(A.) Expert consensus estimates (green dots, with 80% prediction intervals) of the total number of SARS-CoV-2 infections by the end of 2020. Expert predictions aligned with estimates from various computational models during the same time period. Prediction intervals at the 80% level are shown for all estimates except for the Bedford estimate which has a “best guess” prediction interval and a second interval double the size of the first shown as a narrower line [26]. (B.) Expert consensus distributions of the fraction of all infections reported as confirmed cases. In the first three surveys, experts provided a percent of infections that had been confirmed as cases by laboratory test, in the next four (dates in boldface) they directly estimated the total number of infections. Surveys 4-6 asked experts to provide the smallest, most likely, and highest number of total infections, and the last survey asked experts to provide a 10th, 50th, and 90th percentile.

Over the course of seven surveys from March 2^nd^ to April 27^th^, 2020, the median of expert consensus predictions for the percentage of all SARS-CoV-2 (the virus that causes the COVID-19 illness) infections in the U.S. that had been diagnosed was between 6% and 16%. The median responses were consistent with estimates from computational models generated over the same time span[10]–[12], [26]. As a sensitivity analysis, we asked experts to predict the number of hidden infections in two different ways: early surveys asked experts to predict the percent of confirmed cases, and later surveys asked to predict the total number of infections. The first six surveys asked experts to provide a smallest, most likely, and largest estimate. The last survey, on April 27^th^, asked experts to provide a 10^th^, 50^th^, and 90^th^ percentile. We found that the experts’ median prediction of the fraction of total infections that were confirmed was stable when asked to predict percentages versus direct estimates of the number of infections and when asking experts to provide a smallest, most likely, and largest versus a percentile answer.

At the beginning of each of thirteen consecutive weeks from February 17th to May 11th, experts predicted the number of confirmed cases at the end of the week. For surveys administered from February 17^th^ to April 6^th^, participants specified the smallest, most likely, and largest possible number of cases that would occur by the end of the week, and for surveys administered from April 13^th^ to May 11^th^ they assigned probabilities to ranges where the number of cases could occur. (Fig. 3A). In all but two weeks (surveys on March 15^th^ and April 20^th^), expert consensus predictions were more accurate than an “unskilled forecaster”, a naïve prediction that assigned uniform probability across the range of predicted values from all experts. In all but one week (the survey issued on April 27^th^), the consensus prediction was more accurate than the majority of individual expert predictions (Fig. 3B). All thirteen expert consensus 90% confidence intervals covered the reported number of confirmed cases. The consensus prediction was outperformed by expert’s individual predictions 23% of the time and no experts performed consistently better than the equally weighted consensus. As a result, performance-based weighting to build a consensus did not significantly improve forecast accuracy compared to equal weighting (see Supplement).

**Fig 3.**
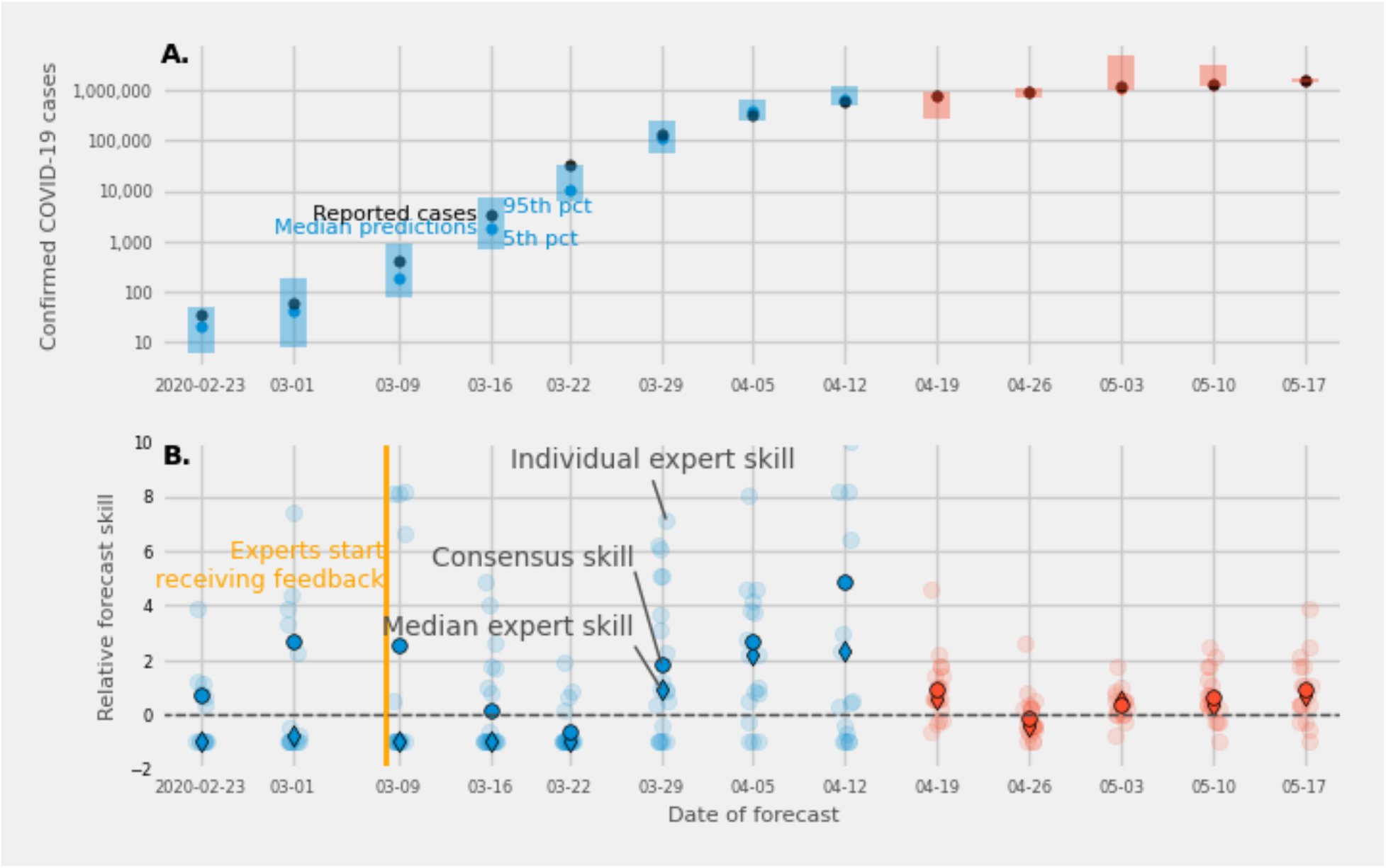
(A.) Expert consensus forecasts of the number of cases to be reported by the end of the week (Sunday, date shown on x-axis), from thirteen surveys administered between February 23 and May 17, 2020. The first eight surveys asked experts to provide smallest, most likely, and largest possible values for the number of confirmed cases (blue bars and dots), and the last five asked experts to assign probabilities to ranges of values for confirmed cases (red bars and dots). Expert forecasts were made on Monday and Tuesday of each week. Light blue and red points represent the median of the expert consensus distribution. Dark points represent the eventually observed value. Prediction intervals at the 90% level are shown in shaded bars. The 90% prediction intervals included the true number of cases on thirteen out of thirteen forecasts. (B.) Relative forecast skill for each expert (light dots), the median expert (dark diamond), and the expert consensus (dark dot), compared with an “unskilled” forecaster (see Methods). Higher relative forecast skill indicates better performance than an “unskilled” forecaster and a zero relative forecast skill represents identical performance with an unskilled forecaster. The expert consensus prediction outperformed an unskilled forecast in all but two surveys. The median expert showed less forecast skill than an unskilled forecaster up until the survey issued on March 23^rd^ (forecasting cases for March 29^th^) and for a survey issued on April 20^th^. Median expert accuracy improved above that of an “unskilled forecaster” (see Methods).

Individual expert predictions did not perform as well as the consensus when experts were asked for a smallest, most likely, and highest number of confirmed cases. The median forecast skill of individual expert predictions was lower than an unskilled forecaster on the first five surveys. The forecasts from individual experts were more accurate in later surveys, and the median accuracy of individual expert predictions was higher than the accuracy of an unskilled forecaster in 7 of the last 8 surveys.

Overall, across the 40 questions about measurable outcomes from February 17^th^ to May 11^th^, a consensus of expert predictions scored better than an unskilled forecaster (Fig. 4A). An expert consensus scored in the top 50^th^ percentile for 31 (78%) questions and had a higher relative forecast skill (relative to an unskilled forecaster) than the median individual expert skill for 36 (90%) questions. Triplet questions, where experts were asked to provide a smallest, most likely, and largest possible outcome, showed the largest improvements between the median expert and consensus forecasts. The consensus was more accurate than all individual experts for 12 out of 21 triplet response questions (Fig. 4B). For percentile and probabilistic categorical questions, the consensus prediction ranked closer to the 50th percentile. An equally weighted consensus is likely to perform better than an individual forecast, but how well a consensus performs may depend on how we ask experts to provide predictions.

**Fig 4.**
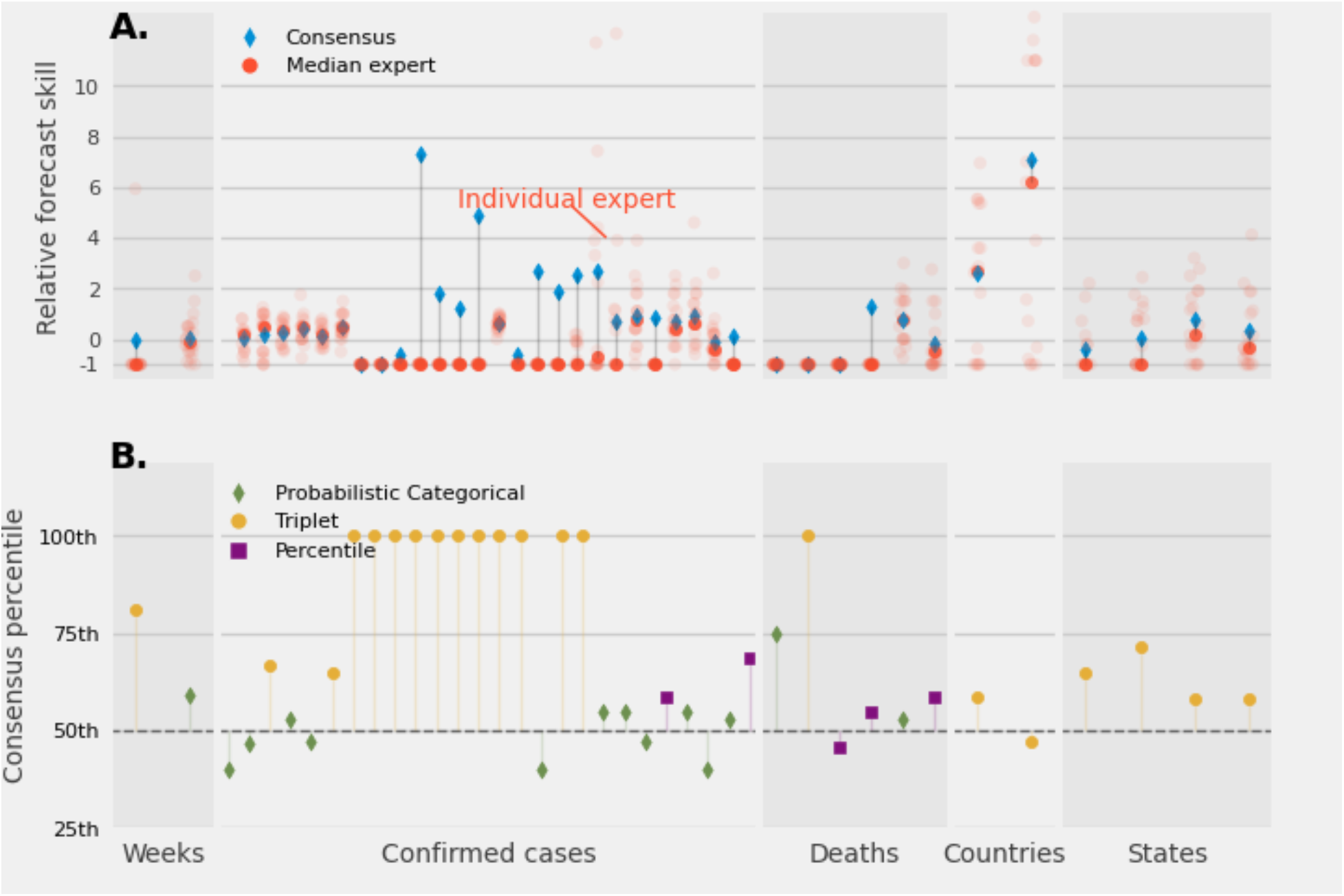
(A.) Relative forecast skill for the consensus prediction (diamond), for individual experts (light circle), and the median (dark circle) of individual expert’s relative forecast skill for 40 questions where the truth could be determined. Predictions are grouped by five different types of forecasting targets: the number of weeks for an event to occur; the number of confirmed cases for one and two weeks ahead and average confirmed cases reported at the state level; number of deaths reported at the state level and short term predictions of the number of deaths for the US; number of countries reporting cases above a specific threshold; and the number of states reporting cases above a specific threshold. (B.) The percentile rank of the consensus prediction compared to individual expert predictions classified by forecasting target and type of answer requested from experts. A diverse range of questions with measurable outcomes was asked. In most cases a consensus distribution led to a more accurate prediction. A consensus most improved questions where experts were asked to provide a triplet answer.

Consensus aggregations of expert judgment during early months of the COVID-19 pandemic provided important and early insights about the trajectory of the emerging outbreak. Since mid-March, when there were less than 100 COVID-19 deaths in the US, expert consensus showed substantial probability of well over 100,000 deaths by the end of 2020. In contrast, early forecasts from a computational model used by the federal government in late March predicted 81,000 deaths and an outbreak that would end by early August [27].

Expert predictions of the US COVID-19 outbreak were well calibrated overall but erred on being too optimistic. Expert predictions of confirmed cases included the true number of cases in their 90% prediction interval for all thirteen predictions. However, accuracy was a challenge. Experts’ predictions in early surveys were smaller than the true number of confirmed cases, but after receiving weekly feedback on their previous predictions (starting on March 9), accuracy substantially improved on triplet questions. The type of answer experts were asked to provide impacted individual accuracy and the variability in individual scores. Experts particularly struggled with triplet response questions.

An expert consensus is a flexible model that can answer critical public health questions before computational models have enough data and validation to be reliable. In particular, an expert consensus model has two key advantages over computational models. First, an expert model has relatively low overhead to develop and can be deployed at the onset of an outbreak. Experts made predictions starting in mid-February before any computational models were available. Second, a survey framework allowed expert predictions to be tailored on-the-fly, to maximize value for public health decision makers.

However, an expert consensus model suffers from issues of scalability. Every individual forecast elicited from an expert requires minutes of human time. Because of experts’ limited time, surveys must focus on a short list of impactful questions. Another potential disadvantage of an expert model is the bias introduced by human judgment. Though the assumptions built into computational models are explicitly specified, experts’ predictive processes are more opaque. A robust forecasting platform, allowing experts to communicate with one another about the reasoning behind their forecasts and interactions between subject matter experts and trained forecasters may lead to more accurate predictions.

An expert judgment model can act as an important component of rapid response and as a first-step forecast for global catastrophes like an outbreak, especially while domain-specific computational models are still being trained on sparse early data. Experts’ ability to synthesize diverse sources of information gives them a unique, complementary perspective to model-driven forecasts that are not able to assimilate information or data outside of the domain of a specific, prescribed computational framework. During the evolving global catastrophe of the COVID-19 pandemic, an expert judgment model provided rapid and well calibrated forecasts that were responsive to changing public health needs.

## Data Availability

Data is available at https://github.com/tomcm39/COVID19_expert_survey

## Acknowledgments

We wish to thank all the experts who have participated, for offering their time and expertise to help us better understand the COVID-19 outbreak. We also thank Evan L. Ray for comments that improved this work.

## Funding

This work has been supported by the National Institutes of General Medical Sciences (NIGMS, grant number R35GM119582) and the Centers for Disease Control and Prevention (CDC, grant number 1U01IP001122). The content is solely the responsibility of the authors and does not necessarily represent the official views of CDC, NIGMS or the National Institutes of Health.

## Supplementary Materials

### Materials and Methods

#### Data Repository

A publicly available repository with details about questions asked and data on all responses is available under a MIT license at https://github.com/tomcm39/COVID19_expert_survey. This is referred to below as “the data repository.”

#### Recruitment of experts

An expert was defined as a researcher who has spent a substantial amount of time in their professional career designing, building, and/or interpreting models to explain and understand infectious disease dynamics and/or the associated policy implications in human populations.

Experts were recruited by sending an email asking for their participation, and by soliciting participation through online forums for infectious disease modelers.

Experts could participate in our surveys after reading and agreeing to a consent document (Fig. S1). The consent states that after an expert completed two surveys, their name and affiliation would be included in public-facing summaries. The consent form also said that public releases of the data would ensure individual expert responses would remain unidentifiable. A current list of experts can be found in the data repository.

Survey data were collected using the web-based Qualtrics platform (Qualtrics, Seattle, WA) through a link sent via email. A link to the survey was also placed on an online forum of modelers focused on COVID-19, asking a participant to self-identify as an expert and fill out the survey. If the participant was vetted to be an expert by the research team, according to the expert definition above, they were added to the list of those that receive weekly emails. Predictions were collected from experts starting on the Monday of each week and closing on Tuesday of that same week.

The proposed research was deemed not human subjects research by the University of Massachusetts-Amherst Institutional Review Board.

#### Survey methods

We asked experts questions that required different types of predictions:

- categorical questions asked experts to pick one out of two (binary) or many options (categorical)
- probabilistic questions asked experts to assign a probability to two (binary probabilistic) or many options (categorical probabilistic)
- percentile questions asked experts to provide a lower (5^th^ or 10^th^) percentile, median percentile, and upper (90^th^ or 95^th^) percentile
- triplet questions asked experts to report a smallest, most likely, and largest possible value for a forecasting target of interest.

A list of all questions and the type of answer required by survey can be found in the data repository.

We deidentified experts by giving them each a unique “expert-id”. To mask expert identities for the data repository, we assigned a different random number between 0 and 5,000 to each expert for every survey.

Data on true outcomes predicted by experts were collected from several sources (Table S1). For questions that have known, measurable answers, we created a database with the observed answers and the resolution criteria – the method used to define the true answer – that can be found in the data repository.

#### Statistical methods

##### Rounding

When reported in the text, expert predictions in the manuscript were taken to have two significant digits. For example, the number 12 was rounded to 12, 123 was rounded to 120, 1,234 was rounded to 1,200, 12,345 was rounded to 12,000, etc. We felt that rounding like this maintained the relevant precision in estimates for public health practice. Rounding was not performed when calculating scores.

##### Scoring predictions

Expert predictions were scored against true outcomes using the log score. The log score is a proper scoring rule that assigns the log of the probability a forecast placed on the true outcome,

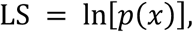

where *p*(*x*)is the forecasted probability assigned to the true value *x*. A log score of 0, indicating the forecast placed a probability of 1 on the truth, is the best score, and a log score of -infinity, indicating the forecast placed a probability of 0 on the truth, is the worst score. Experts’ scores for measurable questions are stored in the data repository.

We used the concept of an “unskilled forecaster” as a benchmark against which to measure predictive performance. For triplet questions, an unskilled forecaster is one who assigns a uniform probability mass to all values between the lowest and the highest predictions any expert proposed for a particular question. An unskilled percentile question assigned to its lower percentile the minimum of all lower percentiles, and to its upper percentile the maximum of all largest percentiles. The unskilled median was the median of all median percentiles. For probabilistic forecasts, an unskilled forecaster assigns 1/C, where C is the number of categories, to each category available as an answer.

A common transformation of the log score is forecast skill, defined as the exponentiated log score. Consensus model performance was reported using the forecast skill of the consensus model divided by the forecast skill of an unskilled forecaster minus one, or the relative difference in forecast skill compared to an unskilled forecaster. A positive difference indicates the model is better informed than an unskilled forecaster.

##### From expert predictions to probabilistic forecasts

Experts made four types of probabilistic predictions: binary, categorical, percentile, and triplet. For binary probabilistic questions, we can define the expert’s prediction of the presence of an event as *p*. Then an expert’s predictive distribution is Bernoulli(*p*). A similar approach can be taken for categorical probabilistic questions, and we define an expert’s predictive distribution over C different choices as a Multinomial(N=1, *p*_1_, *p*_2_,…, *p*_C_) distribution. Percentile questions ask experts to provide three percentiles, a low (5^th^ or 10^th^) percentile, 50^th^ percentile (median), and high (90^th^ or 95^th^) percentile. Specifying three percentiles creates four intervals with probabilities corresponding to the percentiles, for example asking for a 5^th^, 50^th^, and 95^th^ percentile creates four intervals with the following probabilities: 0.05,0.45,0.45, and 0.05 probability, and the probability prescribed to a value *x* was

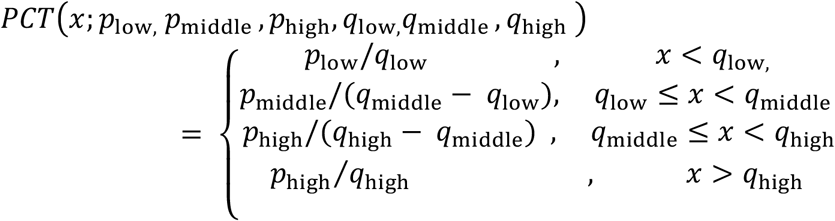

Triangular probability densities (See Fig. S2 for an example) were generated from the smallest (*s*), most likely (*m*), and largest (*l*) answer provided by experts as follows

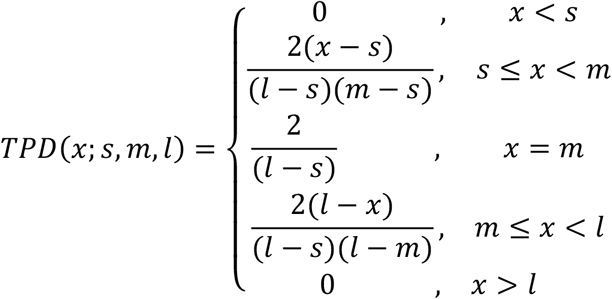

The above TPD specifies an expert-specific probability distribution over a continuous target. To specify a distribution over integers values (*x*_1_,*x*_2_,…*x*_*V*_), we assigned to the value *x*_*i*_ the integral of the continuous TPD from *x*_*i*_up to *x*_*i*+1_. Define the CDF of a TPD distribution as CDF_TPD_[*x*]. Then the discretized TPD was defined as

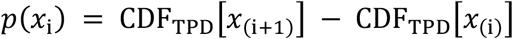

on the values (*x*_1_,*x*_2_,…*x*_*V*_).

##### Aggregation

A consensus distribution for a question (*q*) was created by taking a weighted average of experts’ predictive densities, called a linear pool. We defined a consensus probability distribution for a question *q, f*_*q*_, as

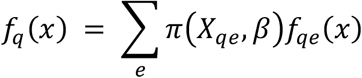

where *X*_*qe*_is a design matrix for each question with one row for each expert and columns for covariates. The entries in the vector *β*are estimated parameters associated with each column (covariate) of *X*_*qe*_, and the matrix product *X*_*qe*_*β*determines how weights are assigned to experts. The function *π*(*X*_*qe*_,*β*) ensures weights are positive and sum to one, and *f*_*qe*_are expert-specific predictive distributions for question *q* and expert *e*. A linear pool is a function of how we assign weights to experts. We chose a specific functional form for weights *π*(*X,β*). The complete model is

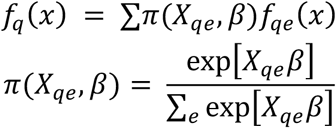

where *π* is the softmax function. An equally-weighted consensus distribution (see Fig. S3. for an example of simulated expert predictions to a triplet question and an equally-weighted consensus) was created by assigning 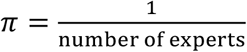 or by assigning *X*_*qe*_= +, an intercept only model. We also examined two additional models. The first model assigned different weights to each expert based on their past performance.

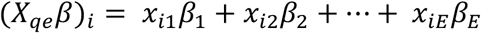

where *β*_*e*_is a weight corresponding to expert *e*, and *x*_*ie*_is equal to 1 when the *i*th observation corresponds to a prediction made by expert *e* and is 0 otherwise.

The third model assigned weights to experts based on expert’s past performance and on the relative entropy of their probabilistic assignment to a question.

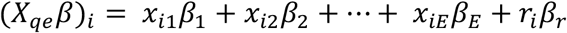

where *r*_*i*_is the relative entropy of the *i*th expert’s prediction and *β*_C_is the corresponding parameter.

Relative entropy is defined as an expert’s entropy of the probability distribution assigned to a question divided by the entropy of an unskilled forecaster. Heuristically, higher entropy indicates that a forecasted distribution is less certain and more uniform in its spreading out of probability across possible values. Entropy is defined for a categorical variable as

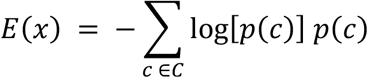

where the variable *x* has *C* categories and for triplet answers was defined as

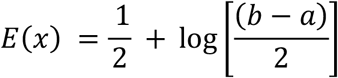

where *a* is the expert’s smallest predicted value and *b* is the largest predicted value. For percentile questions the relative entropy is always one.

Weights for methods of the above form were estimated by optimizing the loglikelihood using PyMC3. The loglikelihood optimized is

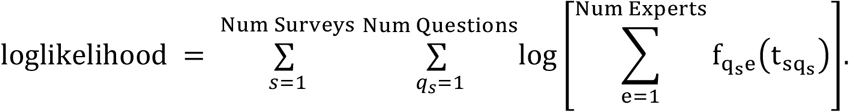

Experts who answered a survey for the first time or had no training data were assigned values equal to an unskilled forecaster, for example a relative entropy of one (i.e. the score of an unskilled forecaster). In later surveys, experts were not required to answer all questions, and in these cases, we assigned them the same value as an unskilled forecaster. These assignments enabled all responses to have observed data with which to calculate weights in a given week.

### Training and testing of weighted linear pools

All consensus distributions distributed to public health officials and the public were built using equally weighted linear pools for all responses.

An equally weighted, expert specific weighting, and expert plus relative entropy weighted ensemble model (Table S2) were retrospectively trained on data from the first survey issued on 2020-02-17 up until the most recent survey issued on May 11th. For each week a survey was issued, past data on expert answers and associated log scores (if the truth would have been available at that time) were available for training and used to estimate weights to assign experts who participated in the present survey. Weights were used to form a consensus distribution and this distribution was scored on the true outcome.

Average and 95% confidence intervals of forecast skill were estimated for each expert, an expert’s average forecast skill over time was estimated, and how forecast skill compared from one survey to the next was estimated (Fig. S4.). The distribution of forecast skill between experts is similar, some experts performing marginally better than others on average. Experts performed similarly over surveys. Performance dipped on surveys issued on 3/16, 3/23, 3/30 likely due to the higher proportion of triplet questions asked. Expert average forecast skill is minimally correlated from one survey to the next and may make it difficult to weight experts based on performance. Performance amongst expert is similar over surveys, through time, and shows a minimal correlation from one survey to the next that is a challenge to assigning consensus weights based on expert performance.

Scores from weighted consensus models were not large enough compared to an equally weighted model to change to a weighted ensemble. The average weight assigned to each of the 41 experts and a table of log scores for measurable questions for each of the three ensemble models is in Figure S5.

Scores across the three consensus models are similar. Because weights assigned to the expert specific and expert specific plus R.E. model are similar, relative entropy does not appear to add information about which experts were more predictive than others. Similar scores between the equal weight and expert specific model could be because an expert’s performance on past survey’s does not suggest future performance on following surveys.

**Fig. S1.**
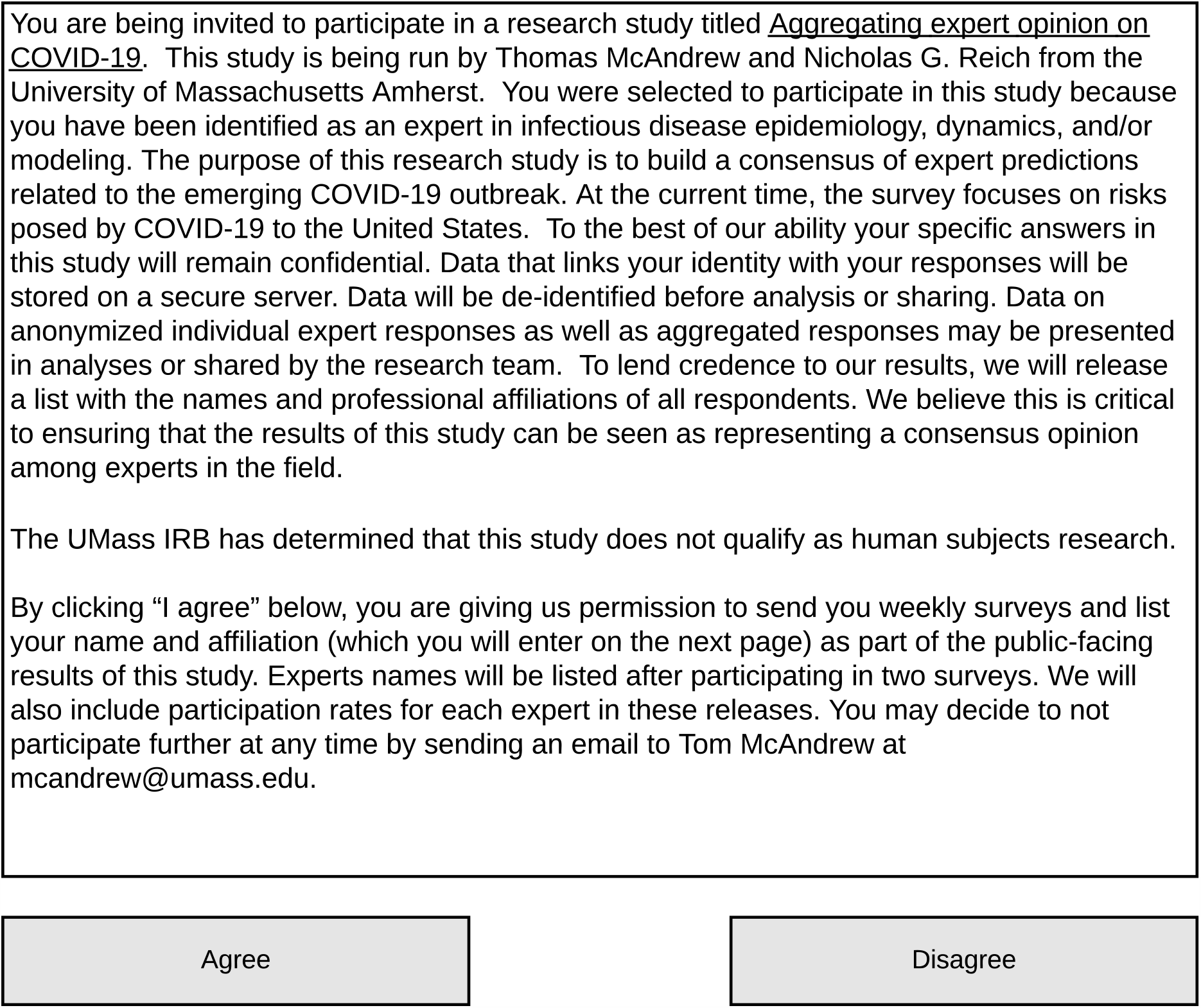
The consent each expert was presented with and had to agree to before taking part in the survey. This document was shown for every survey.

**Fig. S2.**
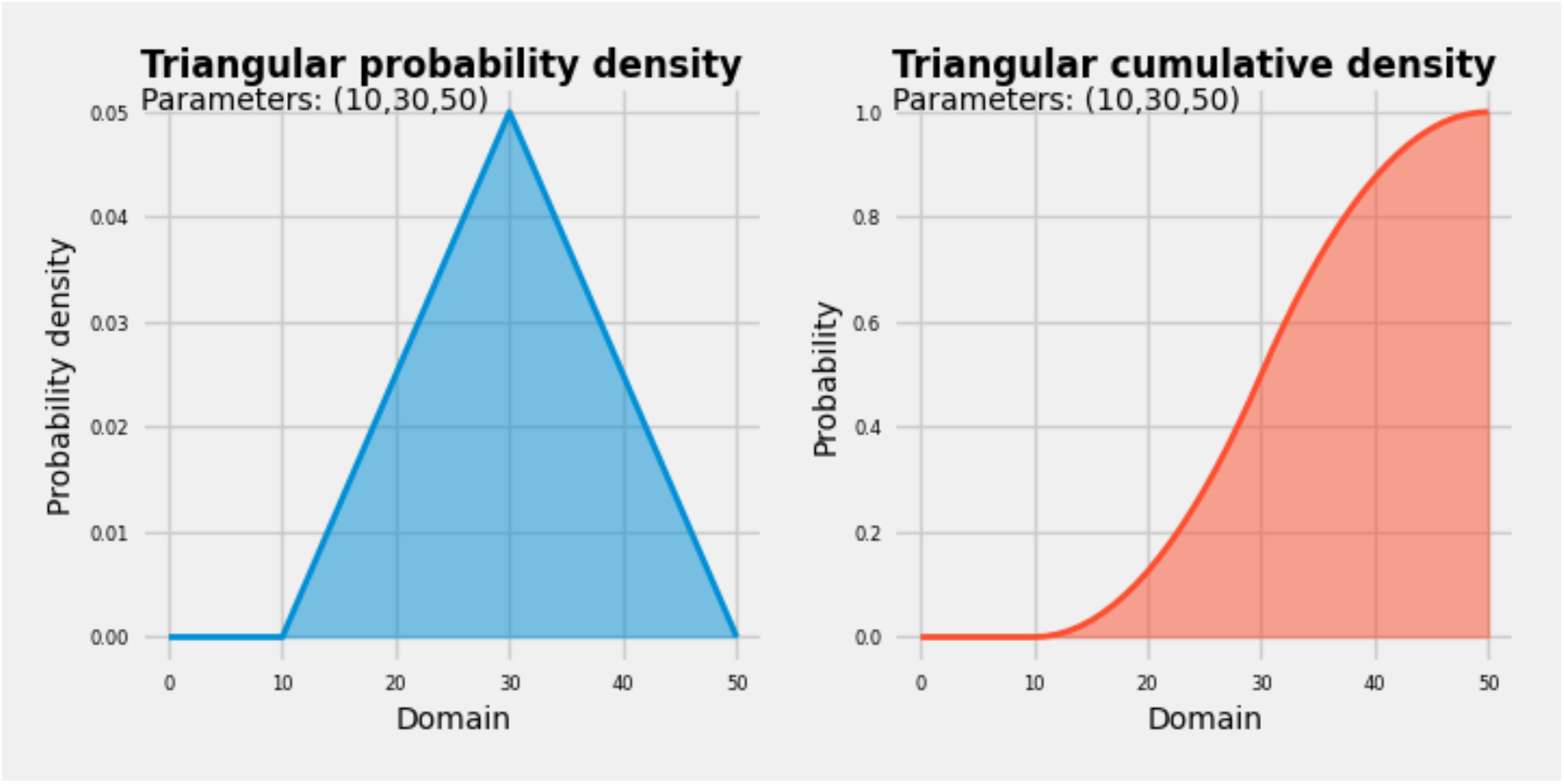
An example of transforming an expert’s triplet answer (smallest: 10, most likely: 30, largest: 50) to a probabilistic distribution.

**Fig. S3.**
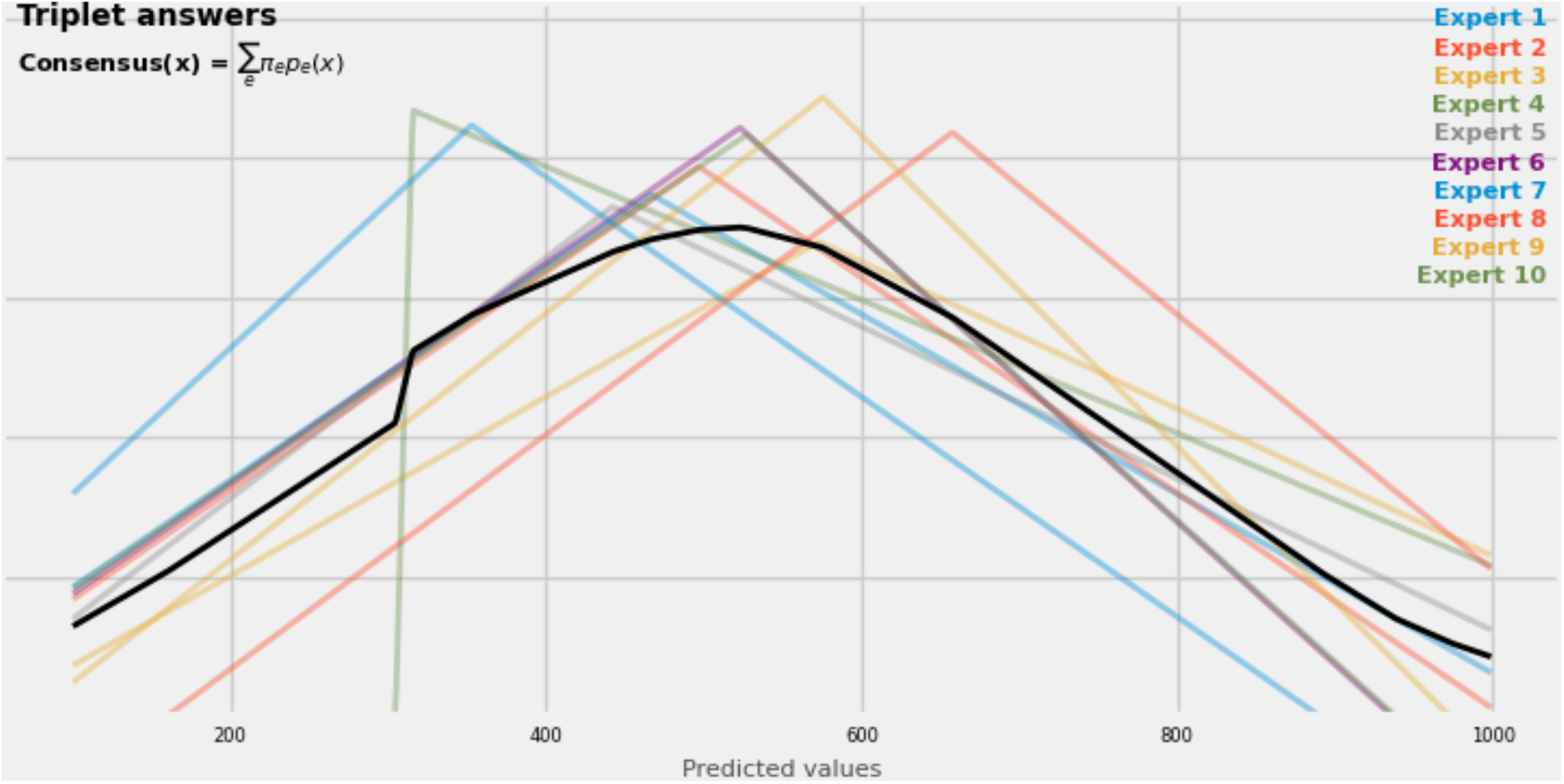
An example of 10 expert answers to a triplet question, their corresponding triangular probability distributions (TPDs), and an equally-weighted consensus distribution built from those TPDs.

**Fig. S4.**
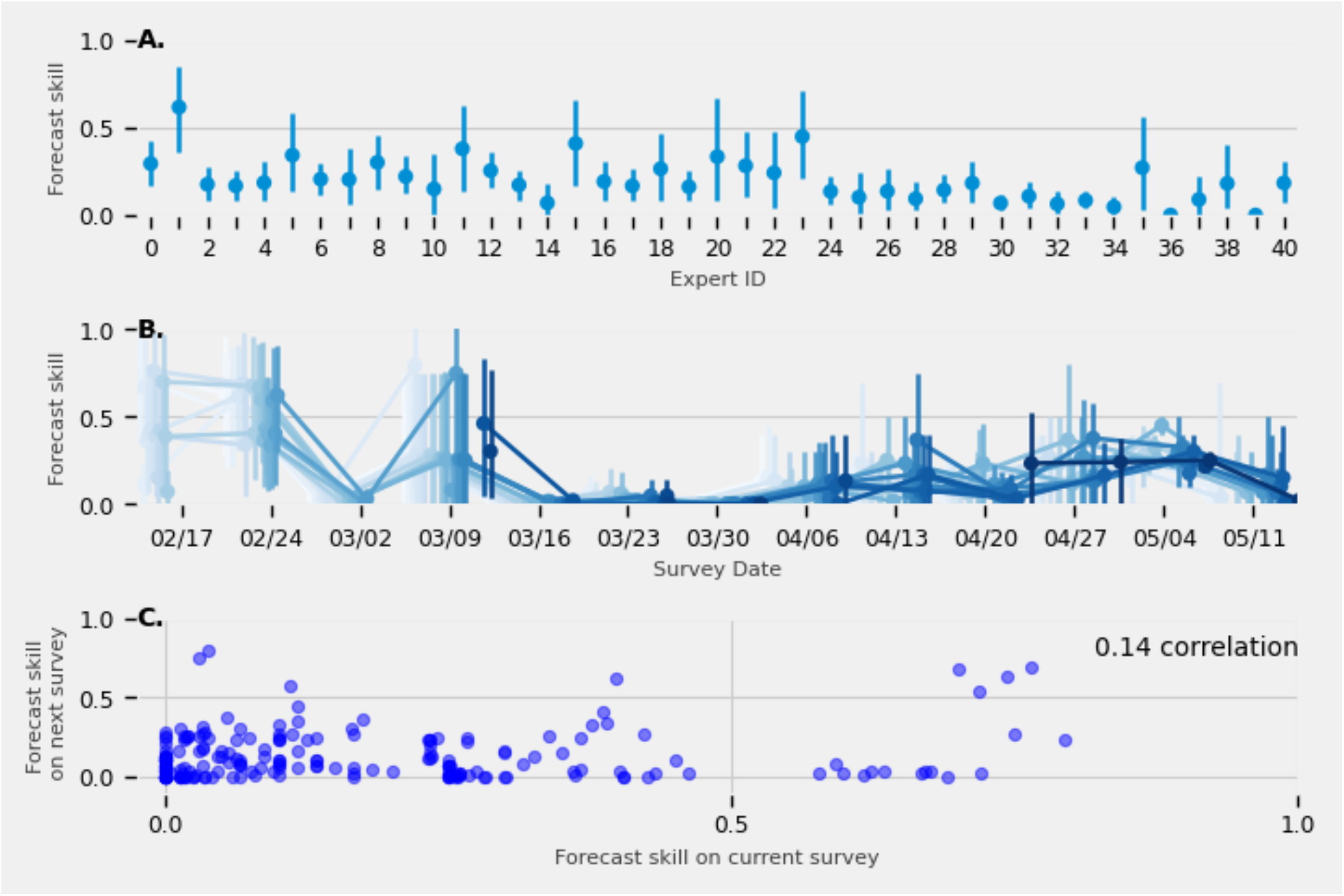
(A.) The average and 95% confidence interval (CI) forecast skill for each expert’s answers over the course of the study from mid-February to mid-May. (B.) For each expert, the average and 95% CI forecast skill across surveys. (C.) For every expert, the average forecast skill for the first survey was plotted against the forecast skill of the second survey they completed, the second against third, up until the average forecast skill for the second-to-last and last survey was plotted. Pearson’s correlation coefficient was computed.

**Fig. S5.**
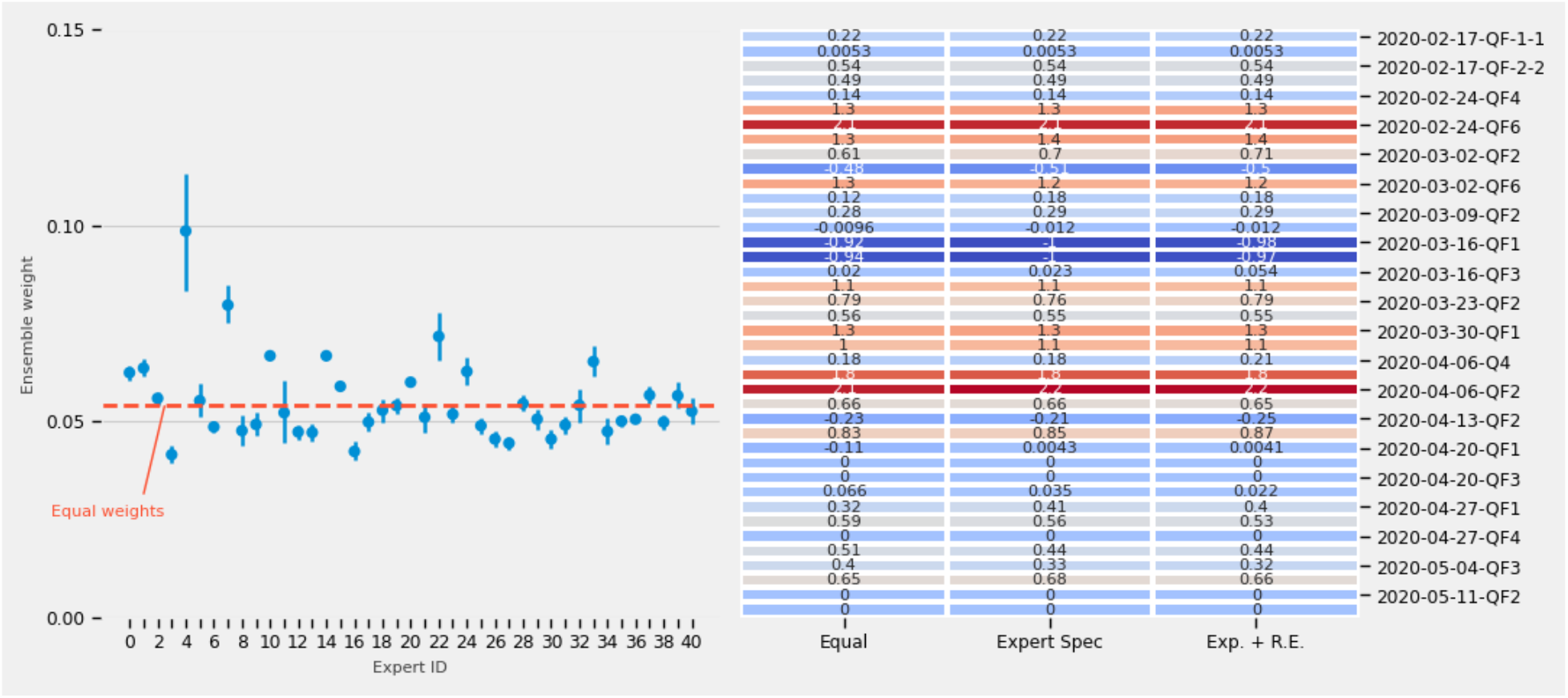
(A.) The average and 95%CI of weights assigned to each expert (from 0-40) over all surveys and questions. The red dotted line denotes the average weight assigned from an equally weighted ensemble. Weights were assigned according to expert’s historical performance (the expert specific model). (B.) The difference between the consensus score and an unskilled forecaster’s score for all measurable questions (40 questions) from three ensemble models: an equally weighted ensemble, an ensemble that assigned weights based on expert’s past scores (Expert Spec), and an ensemble that assigned weights based on expert’s past scores and the relative entropy of expert’s answers to the most recent survey questions (Exp + R.E.). Weights assigned to experts based on past performance are not significantly different than the weights assigned by an equally weighted ensemble. The similarities in weighting translated to similar performance between all three ensembles.

**Table S1.** A listing of survey numbers, the date they were issued, information on expert participation, and the database(s) used to collect ground truth.

| Survey | Date issued | Number of participants | Number of questions asked | Sources for truth |
| --- | --- | --- | --- | --- |
| 1 | 2020-02-17 | 15 | 5 | WHO |
| 2 | 2020-02-24 | 17 | 6 | WHO |
| 3 | 2020-03-02 | 17 | 6 | WHO/CDC |
| 4 | 2020-03-09 | 21 | 6 | CDC |
| 5 | 2020-03-16 | 19 | 6 | CDC/COVIDTracker |
| 6 | 2020-03-24 | 20 | 6 | COVIDTracker |
| 7 | 2020-03-30 | 18 | 7 | COVIDTracker |
| 8 | 2020-04-06 | 20 | 5 | COVIDTracker |
| 9 | 2020-04-13 | 20 | 6 | COVIDTracker |
| 10 | 2020-04-20 | 22 | 7 | COVIDTracker/NYC DOH |

**Table S2.** Model numbers and the covariates used to define the design matrix *X* to weight experts.

| Model Number | Covariates |
| --- | --- |
| Equal | 1 (equal weights) |
| Expert-specific | Expert |
| Expert-specific plus Relative Entropy | Expert , Relative entropy |

## Notes

### Competing Interest Statement

The authors have declared no competing interest.

### Author Declarations

The University of Massachusetts at Amherst Human Research Protection Office determined that this research was Not Human Subjects research, stating "The proposed project does not involve intervention or interaction with individuals OR does not use identifiable private information [45 CFR 46.102(f)(1),(2)]."

